# Genome-based source attribution using a one health *Escherichia coli* isolate collection from 2013-23 in Scotland

**DOI:** 10.1101/2025.11.25.25340865

**Authors:** Antonia Chalka, Louise Crozier, Adriana Vallejo-Trujillo, Vesa Qarkaxhija, Alison Low, Sean McAteer, Kate Templeton, Sue C Tongue, Judith Evans, Geoffrey Foster, Thomas Evans, Charis A Marwick, Ahmed Raza, Benjamen J Parcell, Matthew TG Holden, Tom Mcneilly, Stephen Fitzgerald, Mairi Mitchell, Nuno Silva, Emily Robertshaw-McFarlane, Scott Hamilton, Elizabeth Wells, Clare Hamilton, Eleanor Watson, David Findlay, Julie Bolland, John Redshaw, David Walker, Jane Heywood, Charlotte King, Craig Baker-Austin, Athina Papadopoulou, Andy Powell, Gavin K Paterson, Genever Morgan, Jacqui Mcelhiney, David L Gally

**Affiliations:** Roslin Institute and RDSVS, University of Edinburgh; Food Standards Scotland; The New Royal Infirmary, NHS Lothian, Edinburgh; SRUC Inverness; University of Glasgow; University of Dundee; St Andrews University; Moredun Research Institute; SEPA; CEFAS; University of Liverpool

**Keywords:** **Keywords:** source attribution, *Escherichia coli*, machine learning, one health, food safety, bacteraemia, UTI

## Abstract

Random Forest based source attribution models were developed from a ‘one health’ resource comprising 4,230 high-quality whole genome assemblies from *E. coli*. These were isolated from a wide range of sources, predominantly originating in Scotland, including wastewater, livestock, food, and clinical infections of humans and dogs. Using these models, we derived a probabilistic assignment of *E. coli* isolates from food, shellfish and water samples to potential livestock and human sources of contamination. The incorporation of *E. coli* sequences from wastewater alongside those from human clinical infections, enabled us to capture a wide diversity of human strains in our analyses. The sequence types (STs) of isolates from human bacteraemia and urinary tract infections (UTI) were compared with livestock and food isolates. While only 2.3% of the *E. coli* isolated from food samples in the study were from STs primarily associated with human bacteraemia and UTI, the models found a livestock signal associated with 15% of the human clinical isolates. In the food and private water samples, livestock-human co-attribution of *E. coli* isolates was common and consistent with routine human exposure to specific subsets of livestock *E. coli*, potentially a result of selection during food and water processing. Overall, this research demonstrates the potential value of including source attribution models in national surveillance programmes to understand the transmission of *E. coli* through the agri-food chain and support risk management to protect public health.

**IMPACT STATEMENT:** This research examines the genetic composition of *Escherichia coli* isolated from many different sources including, animals, humans, food, water and wastewater around Scotland. With the public resource generated, machine-learning models were developed to allow the source of an *E. coli*, for example one isolated from food or water, to be predicted from its genome sequence. We show that *E. coli* has genetic content associated with the originating host that allows source tracking using the developed models. Specifically, *E. coli* associated with human UTI and bloodstream infections are acquired predominately form human sources, although 15% of isolates exhibit livestock signals, especially pigs. Food, shellfish and water *E. coli* samples show low association with human clinical strains (∼5%) but over a third of food isolates co-associate to both livestock and human sources supporting food as key pathway to human colonisation. The sequence and associated data provided is a valuable One Health resource the models generated can identify the animal or human source of an *E. coli* isolate. This can help with outbreak tracing and defining the sources of food or water contamination to develop appropriate interventions.

## INTRODUCTION

*Escherichia coli* (*E. coli)* is a ubiquitous commensal in livestock and humans and a key ‘sentinel’ bacterial species used for monitoring faecal contamination in water, assessing food hygiene and for qualifying and quantifying antimicrobial resistance (AMR)[1]. Therefore, *E. coli* can be part of a ‘one sample many analyses’ approach to One Health[2]. The ease and frequency with which *E. coli* can be isolated from many hosts and environments, including humans, livestock, water, soil and food vehicles[3,4], means it is exploited to try and elucidate transmission routes between niches for both the microbe and the genes that it carries, especially those encoding antimicrobial resistance[5–7].

In humans, *E. coli* is the most common cause of human urinary tract infections (UTI)[8,9] and one of the most common causes of lethal bacterial sepsis from bloodstream infection (bacteraemia)[10,11]. In addition, Shiga toxin-encoding *E. coli* (STEC) continue to pose a threat to human health[12,13] and therefore it is important to understand potential sources and routes of transmission of different *E. coli* sub-types to and from humans to protect public health.

The ability of *E. coli* to persist and thrive in many extremely different habitats is directly related to its genomic diversity. *E. coli* has a large pangenome of which approximately 25% of the gene content is core to the majority of strains[14,15]. This wide intra-species diversity makes source attribution for *E. coli* using whole genome sequencing (WGS) challenging as large datasets are required to identify niche, host-related and/or geographically-related signals.

Various types of genome-based signals (features) can be used to infer strain properties such as the originating source of an isolate. In the last decade, machine learning (ML) has been applied to bacterial genome data to predict phenotypes such as antimicrobial resistance[16–19], metabolic pathways[20], disease severity[21,22], phage infectivity[23] and to attribute the source animal[24–28] or geography[29,30] of an isolate. Accurate source attribution helps to define the potential sources of pathogens; thereby supporting the investigation of outbreaks of human illness, or contamination events e.g. to ascertain the source (human, wild birds, canine or agricultural) of *E. coli* in water, which is currently of particular public interest in the UK. The availability of microbial WGS data from statutory and other clinical, environmental and food monitoring and surveillance programmes also provides a valuable source of information to support government strategy for tackling AMR[31].

The potential to use one health genomics for bacterial foodborne pathogens resulted in a UK Government investment of over 20 million pounds in the <u>PATH</u>ogen <u>S</u>urveillance in <u>A</u>griculture, <u>F</u>ood and the <u>E</u>nvironment (PATH-SAFE) programme[32]. Starting in 2021, this programme was led by the Food Standards Agency (FSA) and included pilot projects and infrastructure development for enabling the sharing of bacterial sequence data sets and allied metadata for public health benefit, as with GenomeTracKR in the USA[33].

Food Standards Scotland (FSS) commissioned a pilot project within PATH-SAFE to assess how source attribution modelling could be used within existing surveillance programmes to support understanding of *E. coli* transmission in Scotland. This manuscript reports on; (1) the ‘one health’ *E. coli* dataset established from ongoing sampling networks from across the country; (2) the generation of ML source attribution models applied to *E. coli* isolated from human clinical infections, food, shellfish and water.

## METHODS

### Isolate Collection G Whole Genome Sequencing (WGS)

Isolates were collected from multiple laboratories in order to sample *E. coli* genomic diversity across Scotland including those involved in regulatory monitoring and food surveillance programmes commissioned by FSS, as well as those generated by research projects. Sources included livestock (poultry, ovine, bovine, swine), wild animals (deer, geese), companion animals (canine UTI), food (human and raw pet), humans (UTI and bacteraemia), the environment, drinking and recreational waters, wastewater) and shellfish, primarily as samplers of bacteria in their local marine environments; with a view to capturing genetic diversity of *E. coli* across Scotland. All WGS was carried out by MicrobesNG (https://microbesng.com/). Either *E. coli* isolates or purified DNA were submitted, and llumina sequencing (paired reads) was carried out as described (https://microbesng.com/documents/methods/). The Nextera XT Library Prep Kit (Illumina, San Diego, USA) was used. Libraries were sequenced on an Illumina NovaSeq 6000 (Illumina, San Diego, USA) using a 250 bp paired end protocol. Raw reads were uploaded to Enterobase[34,35] (https://enterobase.warwick.ac.uk/species/ecoli/search_strains?query=workspace:104476).

Of 3,155 Scottish new *E. coli* isolates generated for the project, 3,142 were successfully sequenced and 2,960 assembled. These were combined with 1,354 existing assemblies (human and canine) that were generated through other projects (Supplementary Figure 1). For analysis, 4,230 assemblies passing EnteroBase QC and with <500 contigs were retained. Information on the sources of the isolates is provided in Figure 1 (results section) with complete details in Supplementary Table 1 and a summary in Supplementary Table 2. Imaging and sub-analysis of the data can be carried out at: https://microreact.org/project/pathsafe1b-geninfo (we have not been easily able to remove two duplicated isolates so starting number is 4,232 on the Microreact website).

**Figure 1.**
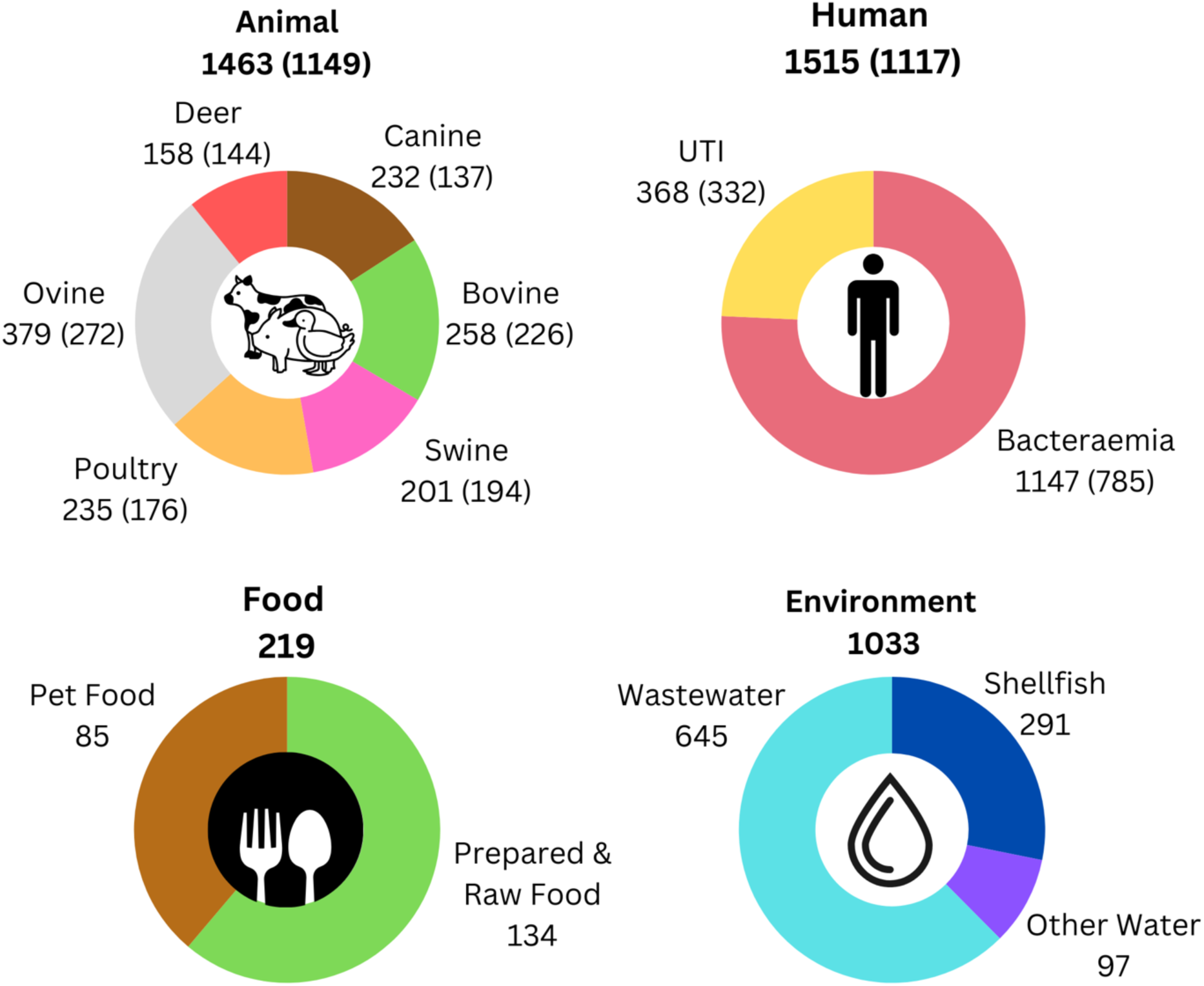
Breakdown of isolate numbers per host in the final 4,230 dataset used for analysis. The numbers in parenthesis represent the isolates used in the nonclonal dataset for model building. A summary of the isolate sources is provided in Supplementary Table 2 and individual isolate metadata in Supplementary Tables 1 & 3. The data can visualised at https://microreact.org/project/pathsafe1b-geninfo.

### Sequence Data Analysis

The workflow for gathering high-quality assemblies, metadata, genome annotation and extraction of features to build and implement ML models was adapted from the methods presented in a previous study[24] and Supplementary Figure 2. High-quality assemblies were annotated with prokka:1.14.5[36], with a nucleotide FASTA file of reviewed *E. coli* proteins from UniProt[37] inputted via the ‘trusted protein file’ option. panaroo:1.5.2[38], run with options: --remove-invalid-genes --merge_paralogs --clean-mode moderate, was used for pan-genome extraction.

For phylogenetic analysis mlst:2.23.0 (Seemann T, mlst - Github https://github.com/tseemann/mlst) was used to define the sequence type (ST). This uses the PubMLST website (https://pubmlst.org/ )[39] . ClermontTyping:24.02[40] for phylogroup identification; snippy:4.6.0[41] to extract core SNPs with strain EC958 as the reference genome[42] with phylogenetic trees built with gubbins:3.2.1[43] and iqtree 2.0.3[44], as well as a binary pangenome presence/absence dendogram. iTOL was used for tree visualisation. SNP distances were calculated using snp-dists:0.8.2[45] to identify epidemiologically linked isolates; those differing by <10 SNPs from the same host were classified as clonal. Of 2,978 high-quality human and animal assemblies, 2,266 non-clonal isolates were used for model training and testing (Supplementary Table 3).

Predictions for both core SNP and pangenome trees were used to generate phylogenetic-prediction models based on 1-5 closest neighbours (Supplementary Tables 4-7). The most commonly occurring host was selected as the prediction. In case of a tie, the host with the shortest average branch length was selected.

### Model Generation

Models were created using the python scientific stack (python:3.11.9[46]; scikit-learn[47]:1.5.1; pandas:2.1.4; numpy:1.26.3;pickleshare:0.7.5). Random Forest was selected for its strong performance on similar datasets[24,28], capacity to model non-linear feature interactions, resistance to overfitting, and ease of feature interpretation. Presence-absence of the pangenome gene clusters were used as input.

Binomial classifiers were built for each host. Model performance was evaluated using Cohen’s Kappa to account for chance agreement. Feature selection was performed using Recursive Feature Elimination (RFE), and models were further refined with hyperparameter tuning. Threshold-moving refined predictions[48] and replaced majority voting with optimized thresholds from ROC and PR curves.

Human attribution models included comparisons with ovine, bovine, swine, poultry, and deer. Two sets of livestock models were created, one where only the livestock isolates were included (‘livestock-only’) and another that included human isolates and labelled as ‘all’ to help remove *E. coli* from the wastewater set that might from non-human sources. To build a ‘human-wastewater’ model aimed at capturing more of the diversity of *E. coli* excreted by humans, wastewater isolates with scores above threshold (defined by ROC/PR curves) in any ’all’ animal model (ovine, bovine, swine, poultry, deer, canine) were excluded. Remaining wastewater isolates, presumed human-associated, were combined with clinical isolates.

To assess model generalizability, reduced human datasets excluded STs with <10 isolates and were sub-sampled to 700, 600, 500, and 400 isolates using StratifiedShuffleSplit[47] (Supplementary Table 8). LGOCV-based on MLST clusters was used to evaluate model robustness to missing phylogenetic data.

Full prediction outputs, including ROC/PR adjustments, are in Supplementary Table 9. To support public-facing interpretation, isolate attribution was visualized using “attribution profiles,” which stack confidence scores from each model, excluding those below threshold. This provides an overview of whether an isolate is attributed to one, multiple, or no hosts. Visualizations were generated in R (R:s4.3.3; Tidyverse:2.0.0; readxl:1.4.3; tidyverse:4.0.5), with full scripts and packages available at GitHub: https://github.com/Antonia-Chalka/stm_ml_pipeline.

## RESULTS

### Diversity of the *E. coli* dataset

From an initial collated set of 4,509 Scottish *E. coli* sequences, a final strain set of 4,230 high-quality assemblies was used in the study which passed quality control thresholds (Figure 1 and Supplementary Tables 1-3, Supplementary Figure 1). This sequence set with metadata (host, time, and, when available, geographical information and antimicrobial phenotyping) can be visualised and analysed at: https://microreact.org/project/pathsafe1b-geninfo.

A whole genome core SNP phylogeny (Figure 2A) was generated for the 4,230 high-quality assemblies and are shown in relation to their isolation source with phylogroups (A-G) and predominant sequence types (STs), according to the Achmann 7 gene MLST scheme. The relative distributions of isolates by source for every ST with over 10 isolates is shown in Figure 2B. Phylogroup with host source is provided in Supplementary Table 10 and ST (inc. subtype), phylogroup and serotype for each isolate in Supplementary Table 11.

**Figure 2.**
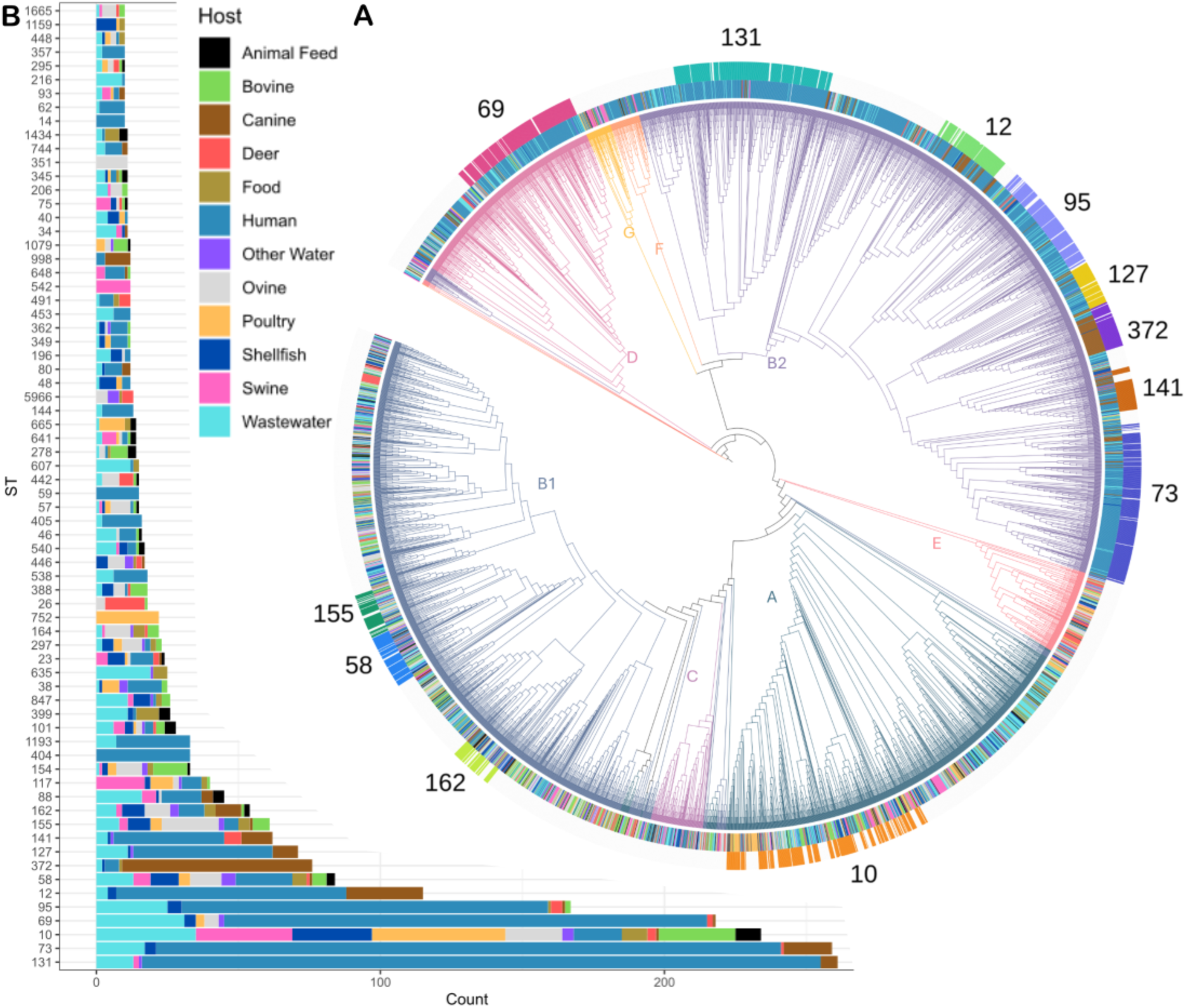
Phylogeny of the E. coli dataset. A) Breakdown of the 7-gene MLST clusters with 10 or more isolates by source. B) Core SNP Tree showing the phylogroup (coloured branches), source (inner coloured ring), and major 7-gene MLST clusters (outer coloured ring). A summary of the isolate sources is provided in Supplementary Tables 10 and individual isolate metadata in Supplementary Tables 11.

The dataset had 881 STs and host specialisation of *E. coli* is apparent within many STs including seven of the top 10 (STs 131, 73, 69, 95, 12, 127, 141) that have predominately human ‘clinical’ isolates (UTI and bacteraemia), ST372 heavily populated with canine clinical strains, ST752 and ST665 with poultry isolates, ST542 for swine isolates, ST351 and ST57 for sheep isolates and deer isolates in ST26. It is also evident that isolates from the various sources are widely distributed and many STs contain isolates from multiple hosts, such as ST10 and ST58, making host assignment based on ST challenging.

Using only clinically-derived human *E. coli* isolates would limit our ability to attribute food and water contamination to human sources using ML. We therefore incorporated wastewater isolates, as many will originate from human faecal matter and will capture some of the human-associated *E. coli* diversity absent from our clinical dataset.

Wastewater isolates were distributed across phylogroups A (33%) and B1 (24%), consistent with commensal strains, with 25% in the more pathogenic B2/D phylogroups. The B2/D isolates clustered within the 172 sequence types (STs) dominated by the human clinical strains, supporting a primarily human source of the wastewater isolates. Most STs enriched from clinical isolates were also found in wastewater, though ST404, ST59, and ST14 were notable exceptions. As a proxy for relative virulence, the proportion of UTI and bacteraemia isolates associated with each main human clinical ST can be divided by their relative proportion in wastewater i.e. an estimate of their general level in wastewater relative to their occurrence in UTI and bacteraemia infections. This ratio has ST131 as the most pathogenic with the following order across the most prevalent STs in the study: ST131 (7.85) > ST73 (5.52) > ST69 (2.33) > ST95 (2.19) > ST10 (0.21). Paired SNP distance analysis highlighted clusters of related isolates across the collection, especially within source groups but also between them, in particular bacteraemia-UTI cases (Supplementary Tables 14 & 15). Specific examples are referred to in later sections.

Most food isolates, including pet food isolates, were evenly split across established commensal phylogroups A and B1. Only a small fraction of the *E. coli* isolated from human food (3/134, 2.2%) and raw pet food isolates (2/85, 2.4%) were found to cluster into the primary human pathogenic STs. These were defined as 172 STs out of the 881 STs in total, for which >50% of the isolates in the ST were from human clinical sources.

FSS undertakes a regulatory monitoring programme to assess the microbiological quality of shellfish growing sites in Scotland[49] which involves the isolation of *E. coli* from bivalve molluscs. 291 *E. coli* isolates from 19 sites across Scotland were sequenced and while the majority were in commensal phylogroups, 19 (6.5%) were in the majority human clinical STs.

### Source attribution models

Based on our published approach[24], random forest models were trained and tested with 2,266 nonclonal assemblies for binary classification i.e. one host vs the rest, with gene cluster presence/absence as features. For machine learning predictions different models were applied: the ‘livestock-only’ set of models for livestock prediction, a human clinical model, and a ‘human-wastewater’ model in which the clinical human *E. coli* isolates were supplemented with a filtered set of wastewater isolates, as defined in Methods.

To benchmark the host attribution models, the accuracy of predictions were compared against other phylogenetic-based methods (Table 1). Specifically, two ‘nearest-neighbour’ phylogenetic models were built, one based on the core SNP tree (Figure 2A) and the other on the pangenome dendogram (<u>Microreact</u>). Models tested 1–5 nearest neighbours, with a single nearest neighbour yielding the best performance. Host prediction based on host ratios within each MLST cluster was also assessed.

**Table 1.**
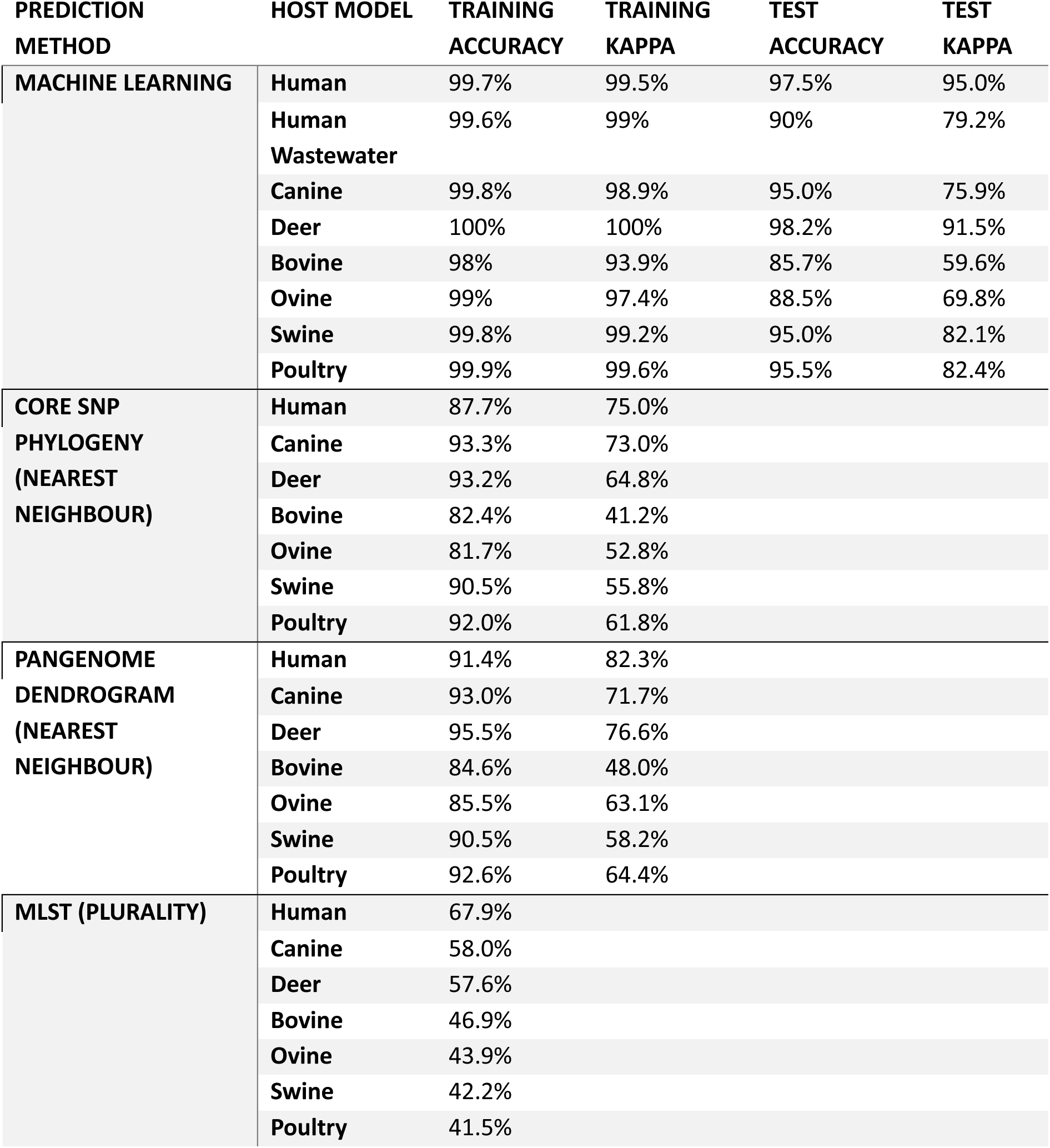
Accuracy metrics across different host attribution models. To evaluate accuracy, both accuracy and kappa statistics were applied. For ML models, accuracy metrics for an unseen test set were generated, and the metrics used are based on PR-corrected thresholds. See Supplementary Table 12 for a full list of the ML metrics and Supplementary Table 13 for the list of isolates used as a test set for each ML model. Phylogenetic predictions were based on the nearest neighbour of a core SNP tree and a pangenome-based dendrogram. Finally, the accuracy of assigning the host to an isolate based on the majority host of the ST it belongs to was investigated.

Phylogeny-based models achieved high raw accuracy (>80%) but showed lower performance on the Kappa metric, which accounts for class imbalance, dropping to 40% for bovine, ∼60% for most animal hosts, and ∼75% for human (Table 1).

Dendrogram-based predictions had higher Kappa than the SNP tree but remained inferior to ML-based models. Model performance was further assessed using an independent test set excluded from training. While accuracy/Kappa declined slightly, ML models remained robust: ∼90% for human and deer, ∼80% for swine and poultry, ∼75% for wastewater and canine, 69% for ovine, and 59% for bovine. Even the lowest-performing ML models outperformed phylogenetic approaches. The reduced accuracy for bovine and ovine models prompted closer examination of misclassifications. Most errors involved mutual misassignment between ovine and bovine isolates, indicating overlap in their genomic signatures (Figure 3A). Livestock models were generally conservative, favouring false negatives over false positives, except in the ovine-bovine cross-over, a preference for non-assignment rather than misclassification. In contrast, the human model showed balanced false positive and negative rates (∼0.02), while the wastewater-human model showed higher false positives (∼0.15) than false negatives (0.07), indicating a tendency to over-assign isolates as human.

**Figure 3.**
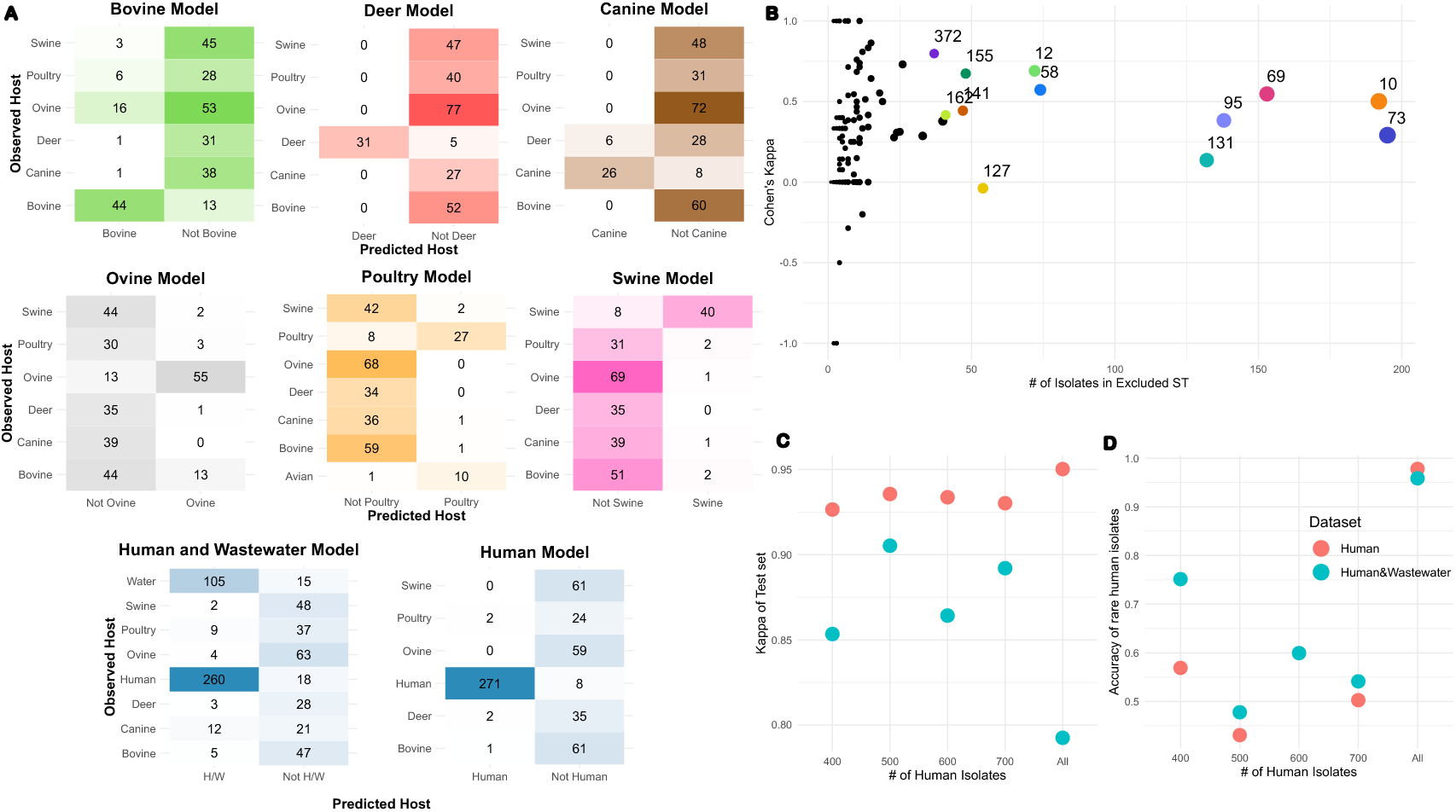
A) Confusion matrices of the ML models’ test sets. As each model was trained as binary classifiers, the predicted hosts are limited to a Host/Not Host classification. A list of test isolates for each model is provided in Supplementary Table 13. B) Leave Group Out Model accuracy. For each model, an ST cluster was set as the test set, with the other ST isolates serving as the training/validation set. Major STs are coloured and labelled. C) Performance of human (red) and human & wastewater models (blue) on complete vs reduced datasets. Model performance measured by the Kappa of the test set. D) Accuracy of the human isolates present in the STs excluded during dataset reduction. A list of which isolates made up each reduced model is provided in Supplementary Table 8.

known potential limitation of the gene-based source attribution models is a reliance on phylogeny for performance[24]. To test if this was present, models were generated using Leave-Group-Out-Cross-Validation based on the 7-gene MLST clusters. As expected, model accuracy varied per excluded clusters but generally led to models with reduced reliability confirming the importance of working within the test population structure (Figure 3B).

One reason for model trends, as defined above, is the imbalanced dataset, in particular the higher number of human and human-wastewater isolates compared to those from other sources. To address this, these groups were sub-sampled while preserving diversity. Kappa values remained high, though accuracy declined for rare human STs excluded from training (Figure 3C). For assigning isolates to a potential human source, the sub-sampled 600-isolate human-wastewater model was used, which balanced sample size with representative ST diversity (Figure 3D).

### Source attribution of *E. coli* associated with human clinical disease, food, shellfish and water UTI and Bacteraemia Isolates

Out of the 1,515 human clinical *E. coli* isolates, 228 (15.0%) attributed to a livestock host model, often with a human co-attribution. Specifically, 150 (9.9%) isolates attributed to swine and human, and 13 (0.9%) isolates attributed to swine only. 21 (1.4%) isolates attributed to poultry and human, 6 (0.4%) to poultry only. 8 (0.5%) isolates attributed to bovine and human, and 7 (0.5%) to bovine only. 4 (0.3%) isolates attributed deer and human 1 (0.1%) to deer only. 4 (0.4%) isolates attributed to ovine and human. A few isolates were attributed to three hosts, such as 5 (0.3%) isolates that were attributed to swine poultry and human, and 2 (0.1%) isolates that attributed to bovine and swine and human. Some isolates were co-attributed to livestock only, such as 5 (0.3%) isolates that attributed to swine and poultry, 1 (0.1%) isolate that attributed to bovine and swine. So, while most human UTI and bloodstream infection strains appeared quite human-specific, a small subset have genetic content associated with livestock strains, particular pigs (N= 176, 11.6%, when combining all associations above).

#### Food Samples

In this study, 219 *E. coli* isolates were sequenced from various food sources, including raw meat, raw pet food, salads, dairy, and ready-to-eat items. Attribution results (Figure 4) are based on the livestock and reduced human-wastewater models. Among the 134 human food samples (Figure 4A), 100 (74.6%) were confidently assigned to a livestock source. For raw pet food (n=85), (Figure 4B), 40 (47.0%) of the confidently attributed isolates were found to match the expected production animal. Concordance was reasonable for beef, poultry, and lamb products, despite the ovine-bovine overlap. In contrast, *E. coli* from venison often misassigned to other species.

**Figure 4.**
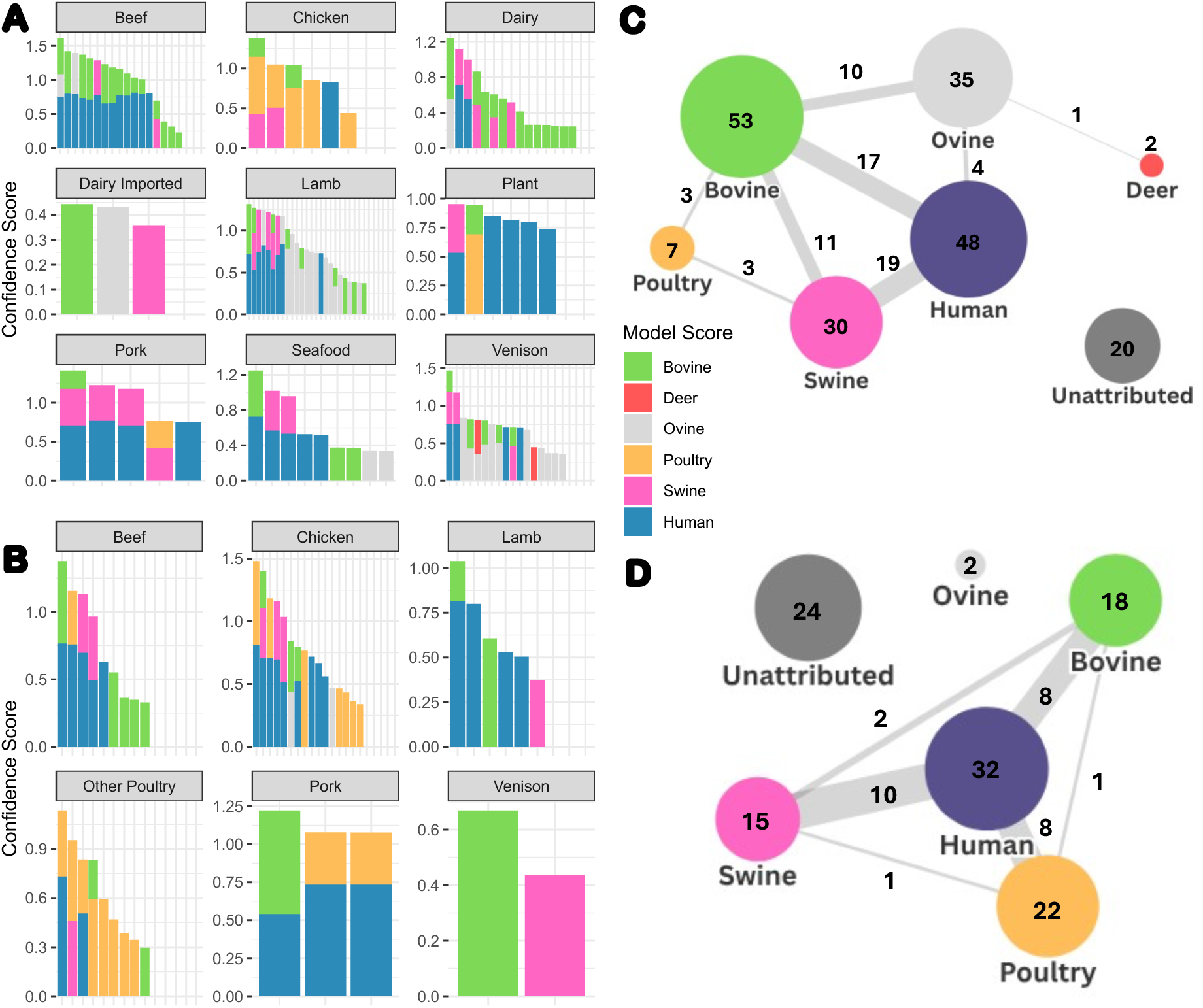
Food isolate attribution. The models used for human attribution are the human & wastewater reduced 600 models highlighted earlier (Figure 3D). Only confidence scores above the threshold for attribution are shown for each sample. The full attributed food set is in Supplementary Figure 3. For bar graphs, each bar represents an individual isolate’s attribution profile. A) Attribution of prepared and raw foods intended for human consumption. B) Attribution of raw pet foods. C) Network chart of human food isolate attribution. Each node (circles) represents the set of isolates assigned to a host or unassigned. The connecting lines represent the number of isolates that have multiple assignments across the two hosts. D) Network chart of pet food isolate attribution.

Of the 219 food-derived *E. coli* isolates (human and pet food combined), 94 (42.9%) assigned only to livestock, while 80 (36.5%) showed significant human attribution from which 58 (26.5%) co-assigned to a livestock source, while 22 (10.0%) were assigned only to the human-wastewater model. These were mostly non-pathogenic phylogroups and STs, although there were some exceptions; (1) an isolate from a chicken sandwich which assigned only to a human source from the models and this closely matched a clinical bloodstream isolate (3 SNPs apart); (2) an isolate from a venison burger belonging to ST95, an important pathogen-associated sequence type. It is worth noting that 20% of the food isolates lacked any clear human or livestock attribution. The general relationships are summarised in Figure 4C for human food and Figure 4D for raw pet food with </=5 SNP relationships summarised in Supplementary Table 15 and detailed in Supplementary Table 14.

#### Shellfish samples

The shellfish samples used in this study were collected from nineteen different sites across Scotland and 291 sequences analysed using the source attribution models (Figure 5).

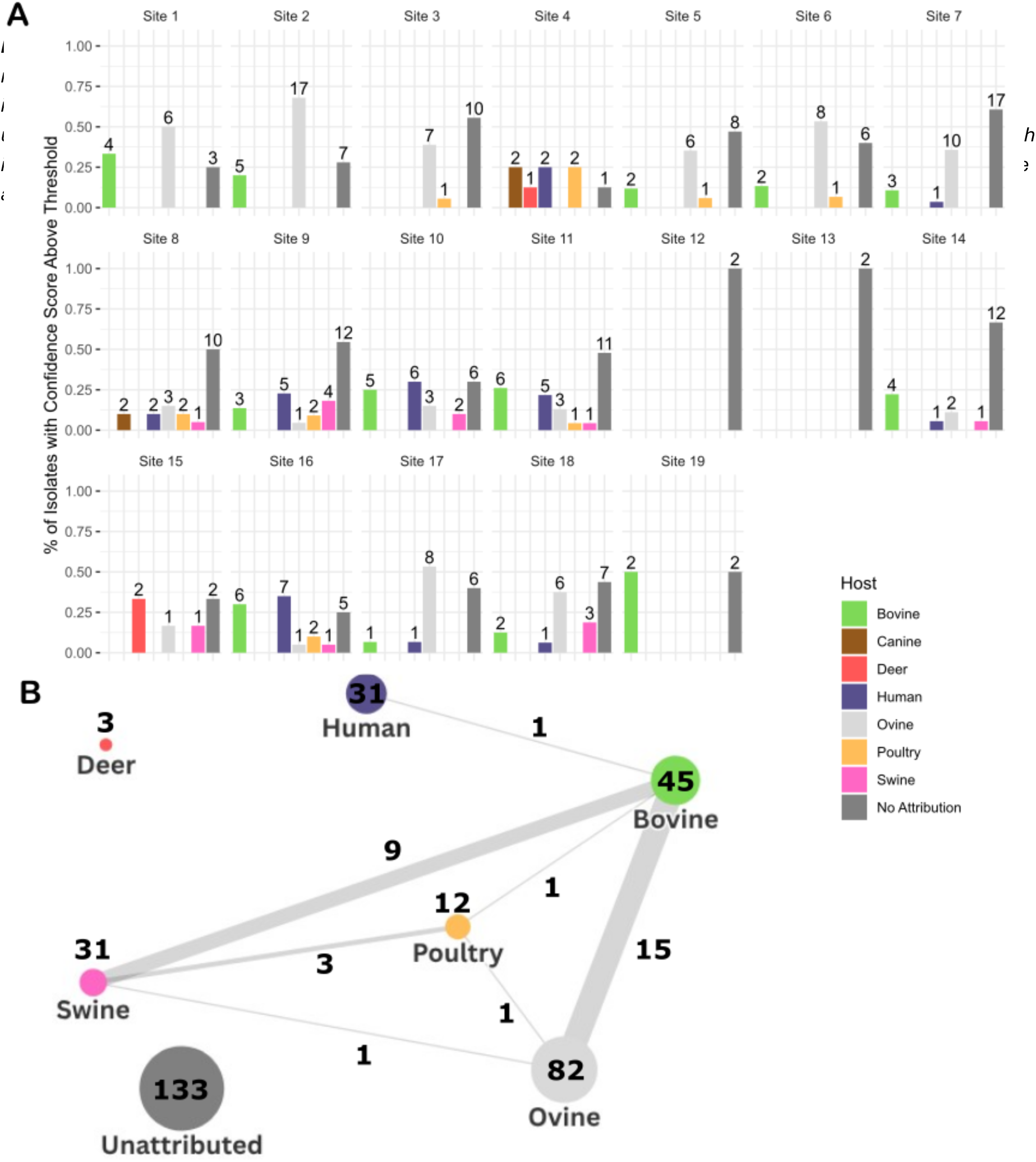

The findings indicate that attribution varied markedly with respect to shellfish sampling location. Twelve of the nineteen sites had a predominant ruminant attribution (ovine and bovine), which suggests that these sites are likely to be located in proximity to agricultural environments. Out of the 291 samples, 30 (10.3%) attributed exclusively to a human source; for three sites this was >20% of the samples; indicating that these sites may be at greater risk from wastewater discharge or direct human contamination. Other non-livestock sources of *E. coli*, such as canine (2 sites), were detected (Figure 5A). While many of the shellfish isolates belonged in rarer STs and will require a larger test collection set to ascribe the specific source of such isolates with higher confidence, there were 14 (4.8%) isolates belonging to the main human pathogenic lineage STs. It was notable that from the shellfish samples there was virtually no co-attribution (one isolate only) of the human assigned isolates with livestock sources. This contrasts with the high level of co-attribution demonstrated for human food and raw pet food isolates (Figure 5B compared to Figures 4C-D).

### Water samples

Our analysis also included 97 *E. coli* isolates from other water samples. These originated mostly 73/97 (80.2%) from private water supplies, with the remainder comprising a mixture of environmental and recreational sources. Of the 73 private water supply isolates, 17 (23.2%) assigned to human only, 10 (13.7%) to both human & livestock, and 56 (76.8%) assigned only to livestock hosts. The network chart for private water isolates is provided as Supplementary Figure 4.

## DISCUSSION

This study utilized national surveillance and monitoring programmes to compile a novel dataset of 4,230 *E. coli* genome assemblies from a range of One Health sources. A key application for the dataset was the development of host attribution models for *E. coli* using machine learning (ML) as a tool to support understanding of transmission pathways and potential sources of contamination for food and water samples. Building on prior work predicting isolate source and infection risk[24,27,28], this study demonstrated that ML models, using differential gene features, outperformed phylogeny-based clustering in accuracy. As defined previously, model accuracy was heavily dependent on staying within the population diversity of the training set[24].

Specific models were developed, including one that combined sequence data from the human clinical isolates and a subset of wastewater isolates to capture a wider diversity of *E. coli* for attributing human/wastewater-based contamination.

While most human clinical (UTI and bacteraemia) isolates belonged to dominant human STs (131, 73, 69 and 95)[9], the source attribution models indicated 16% may be linked to a livestock origin, with the highest attribution to pigs (11.6%). A study in the USA, based on mobile genetic elements[50], estimated that 8% of clinical *E. coli* (mainly UTI in their study) may have a meat-animal source. It is appreciated the ML approach lacks the precision of SNP-based sub-clustering to identify, for example, host switching events and emergence of host specific sub-clusters. However, individual strains of *E. coli* can colonise multiple hosts and a probabilistic approach, sometimes with multiple assignments, is a valid additional analysis to attribute sources of infections, even if the timescales for attribution are unclear.

Potentially in relation to the above, a key finding was that many human and pet food isolates, even though attributing to a logical livestock source (for example 70% of beef products were attributed to cattle) also had a significant human co-attribution (80/219, 36.5% of the combined food isolates, Figure 4C/D). This contrasts with the livestock attributed isolates from shellfish production sites which had very little human co-attribution (1/291, 0.3%, Figure 5B). The co-attribution of isolates from private water supplies was 10/73 (13.7%, Supplementary Figure 4). This would indicate that livestock *E. coli* isolated from food (and to a lesser extent private water) do not represent the wider diversity of *E. coli* excreted by livestock, with one explanation being that these isolates are specific subsets that survive food and water processing and then colonise humans as a result of routine exposure through consumption.

Assigning human origin to food and water isolates was challenging, but inclusion of wastewater isolates significantly enhanced the process. While excluding livestock isolates, based on our livestock models, from the combined human clinical–reduced wastewater model introduced some circularity, it enabled representation of many human-associated *E. coli* not linked to disease. 12% of the food isolates assigned uniquely (no other assignment) to a human source using the reduced human clinical and wastewater model, with a quarter of these (3% of food isolates) matched to major human clinical STs (UTI/bacteraemia). These isolates could be examples of human contamination, directly or indirectly, in the food chain with the potential to result in clinical illness.

Our study found that ∼25% of wastewater isolates belonged to the major clinical STs. This indicates their common carriage and faecal excretion by the general population. It was interesting to note that ST131 and ST73 showed the greatest disparity between their proportions in wastewater and proportional associations with clinical infections (UTI and bacteraemia). This ratio supports the idea that these particular STs have higher pathogenic potential rather than being present at higher overall frequencies in the human GIT. More research is needed to compare levels and types of *E. coli* routinely excreted in faeces and urine from humans to understand how well wastewater samples provide a proxy for this.

The study included samples from ranging ‘wild deer’ and there were individual examples of specific human origin pathogenic ST types (ST95, 73 and 69) being recovered from deer, potentially indicating their exposure to human contamination in the environment. In a similar way, shellfish *E. coli* levels are monitored as a signal of seawater contamination in the production area and our models have then allowed source attribution of shellfish isolates from different cultivation sites around Scotland. 13/19 sites had mainly ruminant-associated *E. coli* consistent with agricultural run-off into the shellfish production site. However, *E. coli* isolates from a few of the sites had low livestock but high relative human attribution; in total 10% of the shellfish isolates assigned to only the human-wastewater model. There were several examples of shellfish isolates which were 0 and 1 SNP apart from those found in wastewater, indicating that the sites from which these samples were taken had been subject to contamination via wastewater. These findings highlight the potential of the source attribution models to provide information that could assist authorities and businesses in developing mitigation strategies which can manage the risks of contamination.

Further validation of our attribution models against existing microbial source tracking methods would be useful to assess the value of integrating *E. coli* WGS into existing programmes for the monitoring and management of water that is intended for consumption, recreational and food production purposes. To further develop the models, as well as direct human gut isolate sequences, broader sampling is needed from sources like rodents, wild birds, and natural environments including soil, water and plants. These will likely fill gaps where we currently have no assignment over our set thresholds.

## CONCLUSION

This study demonstrates the potential value of ‘multi-source’ isolates for the development of ML attribution models and how these models can be strengthened through the integration of wastewater isolates to increase the diversity of human associated strains available for analysis. Although it only provides a snapshot of *E. coli* diversity, the dataset signposts strain transfer across humans, animals, food, and water. It also provides a valuable resource for interrogating AMR allele flow in a one health network and research is ongoing with this analysis. These attribution models should also assist in outbreak investigations, for example ongoing research is examining the utility of the *E. coli* attribution models to understand sources and threat from different serotypes of Shiga toxin producing *E. coli* (STEC) in the UK. Overall, the study underlines the need to optimise One Health surveillance programmes towards identification of interventions that can protect public health.

## Data Availability

All data produced in the present work are contained in the manuscript or via the links provided below

https://microreact.org/project/pathsafe1b-geninfo

https://enterobase.warwick.ac.uk/species/ecoli/search_strains?query=workspace:104476

https://github.com/Antonia-Chalka/stm_ml_pipeline

## DATA SUMMARY

The genomes and associated data can be viewed and interacted with at: https://microreact.org/project/pathsafe1b-geninfo

The raw reads are available at: https://enterobase.warwick.ac.uk/species/ecoli/search_strains?query=workspace:104476

Full scripts and packages available at GitHub: https://github.com/Antonia-Chalka/stm_ml_pipeline.

All data other than sequence data for the study are available in Supplementary Tables 1-14.

**We confirm all supporting data, code and protocols have been provided within the article or through supplementary data files.** ☒

## Ethical statement

The majority of the samples were obtained as part of routine surveillance or did not require ethical permission. The exception were clinical isolates obtained under Lothian SR1094, 15/ES/0094.

