## Supplementary Figures for "Genome-based source attribution using a one health *Escherichia coli* isolate collection from 2013-23 in Scotland"

This supplementary material is hosted by *Eurosurveillance* as supporting information alongside the article [Genome-based source attribution using a one health *Escherichia coli* isolate collection from 2013-23 in Scotland], on behalf of the authors, who remain responsible for the accuracy and appropriateness of the content. The same standards for ethics, copyright, attributions and permissions as for the article apply. Supplements are not edited by *Eurosurveillance* and the journal is not responsible for the maintenance of any links or email addresses provided therein.

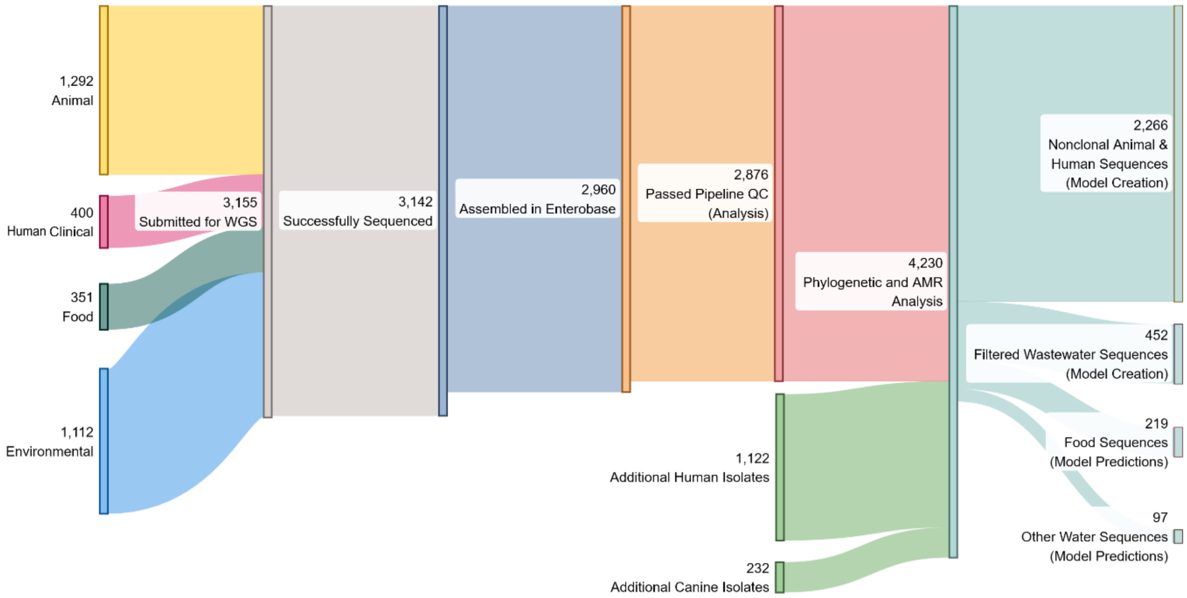


**Supplementary Figure 1**: Outline of sequenced and retrieved isolates. Not all collected isolates led to a usable sequence, due to sampling or assembly QC metrics.

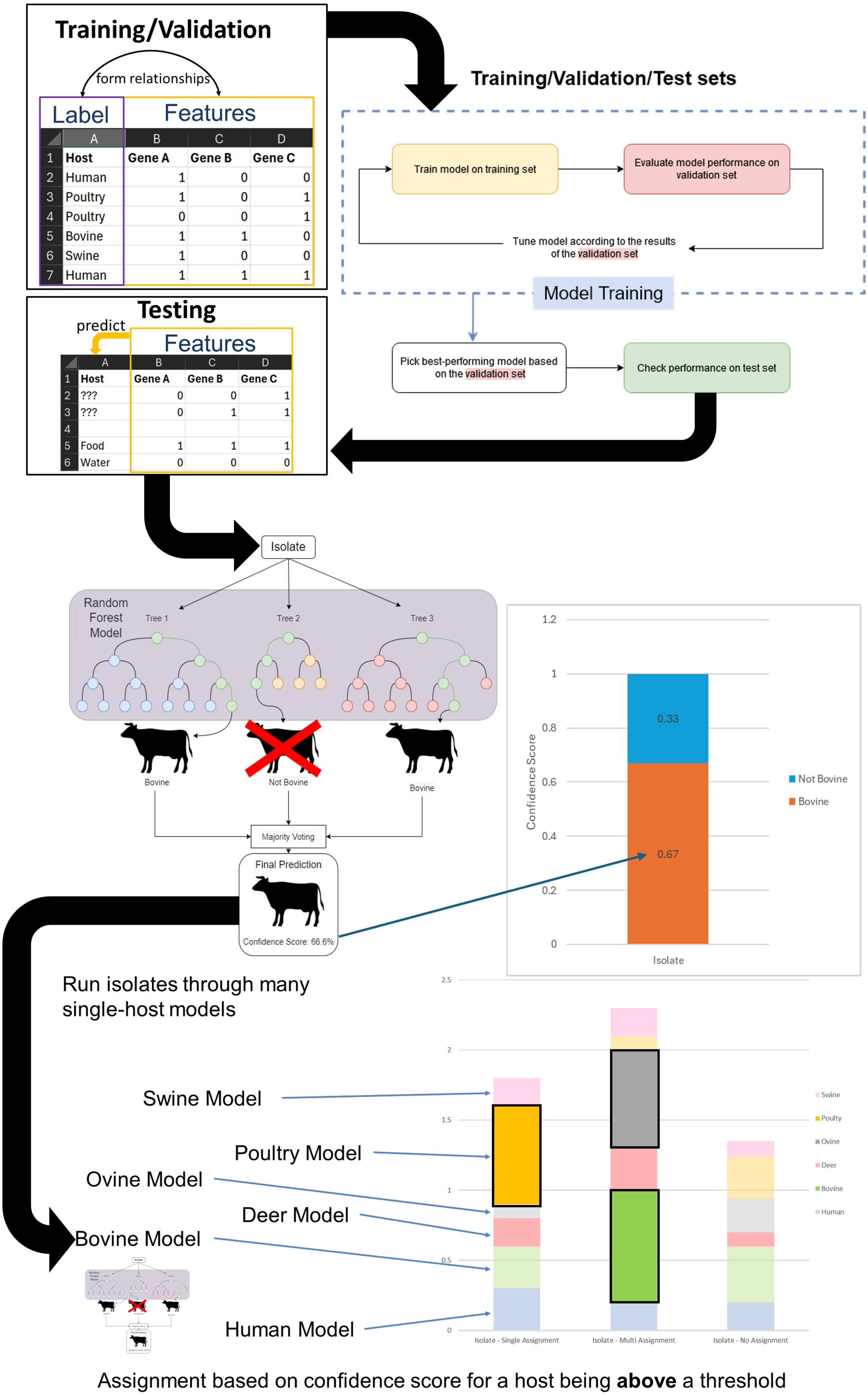


**Supplementary Figure 2**: Outline of attribution profile creation for individual isolates. The inputted dataset are host labels (converted to binary Host/Not Host for each model), and gene presence/absence as the features the models will use to make predictions. A Random Forest model was created based on a training/validation set and its performance is checked on an unseen test set. The score for each host is based on the confidence score of the model, which is the % of votes the host receives across all the individual decision trees that make up that host's Random Forest model. The score for each host is collected and stacked on each other. Scores below the threshold for attribution (which are corrected using Precision Recall curves) are removed from the chart.

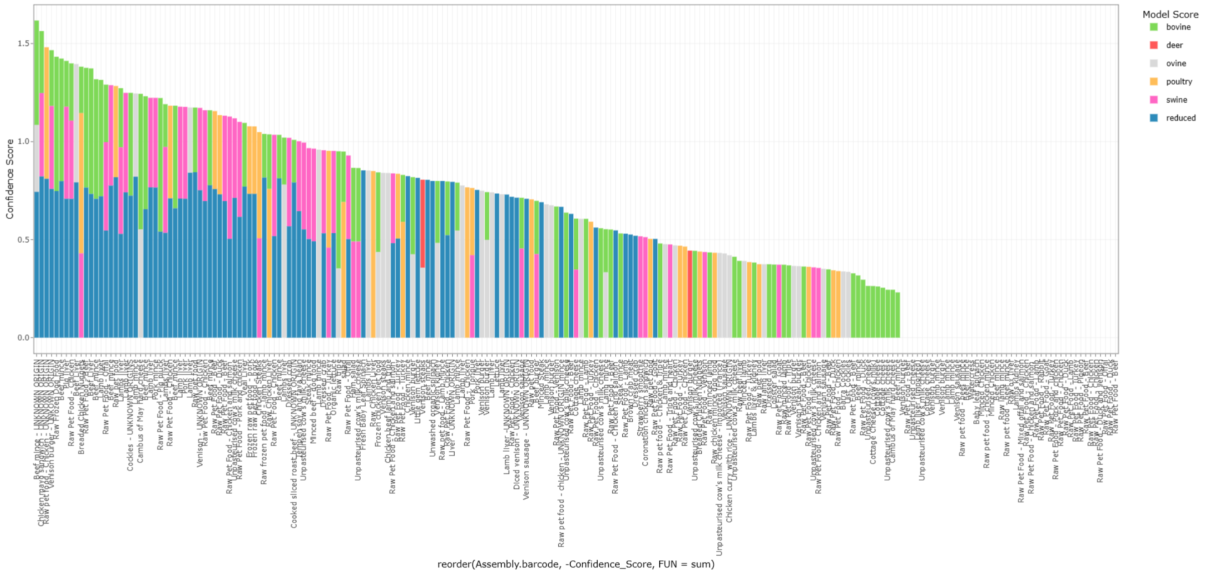


**Supplementary Figure 3**: Attribution of all food isolates across our dataset.

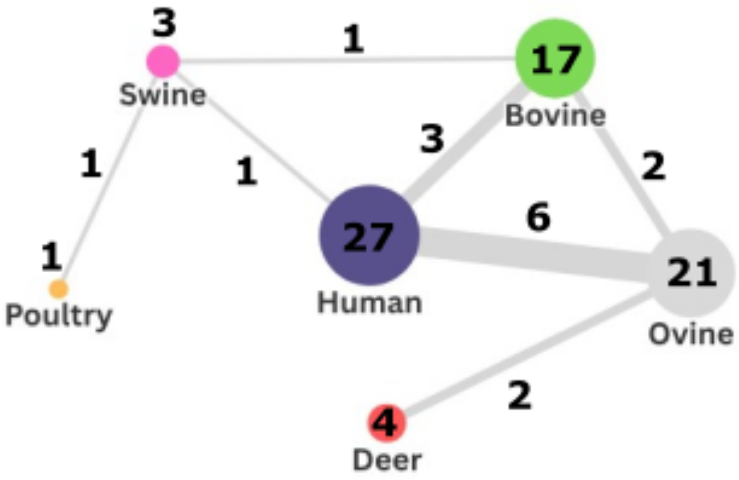


**Supplementary Figure 4**: Network chart of private water attribution. Each node (circles) represents the set of isolates assigned to a host/unassigned. The connecting lines represent the number of isolates that have multiple assignments across the two hosts.
